# SARS-CoV-2: Proof of recombination between strains and emergence of possibly more virulent ones

**DOI:** 10.1101/2020.11.11.20229765

**Authors:** Dania Haddad, Sumi Elsa John, Anwar Mohammad, Maha M Hammad, Prashantha Hebbar, Arshad Channanath, Rasheeba Nizam, Sarah Al-Qabandi, Ashraf Al Madhoun, Abdullah Alshukry, Hamad Ali, Thangavel Alphonse Thanaraj, Fahd Al-Mulla

**Affiliations:** Department of Genetics and Bioinformatics, Dasman Diabetes Institute, Kuwait; Department of Biochemistry and Molecular Biology, Dasman Diabetes Institute, Kuwait; Public Health Laboratory, Ministry of Health, Kuwait; Department of Otolaryngology & Head and Neck Surgery, Jaber Al-Ahmad Hospital, Ministry of Health, Kuwait; Department of Medical Laboratory Sciences, Faculty of Allied Health Sciences, Health Sciences Center, Kuwait University, Kuwait

## Abstract

COVID-19 is challenging healthcare preparedness, world economies, and livelihoods. The infection and death rates associated with this pandemic are strikingly variable in different countries. To elucidate this discrepancy, we analyzed 2431 early spread SARS-CoV-2 sequences from GISAID. We estimated continental-wise admixture proportions, assessed haplotype block estimation, and tested for the presence or absence of strains recombination. Herein, we identified 1010 unique missense mutations and seven different SARS-CoV-2 clusters. In samples from Asia, a small haplotype block was identified; whereas, samples from Europe and North America harbored large and different haplotype blocks with nonsynonymous variants. Variant frequency and linkage disequilibrium varied among continents, especially in North America. Recombination between different strains was only observed in North American and European sequences. Additionally, we structurally modeled the two most common mutations D614G and P314L which suggested that these linked mutations may enhance viral entry and stability. Overall, we propose that COVID-19 virulence may be more severe in Europe and North America due to coinfection with different SARS-CoV-2 strains leading to genomic recombination which might be challenging for current treatment regimens and vaccine development. Furthermore, our study provides a possible explanation for the more severe second wave of COVID-19 that many countries are currently experiencing presented as higher rates of infection and death.

## Introduction

The recent severe acute respiratory syndrome coronavirus-2 (SARS-CoV-2) outbreaks have grievously impacted the world by threatening lives and impeding human activity in a short period. Understanding the factors that govern the severity of a pandemic is of paramount importance to design better surveillance systems and control policies [1]. In the case of COVID-19, three variables play a critical role in its spread and severity in a country: the nature of the pathogen, genetic diversity of the host population, and environmental factors such as public awareness and health measures provided by governments [2].

SARS-CoV-2 spike (S) glycoprotein plays an integral role in the viral transmission virulence [3]. The S protein contains two functional subunits S1 and S2 cleaved by FURIN protease at the host cell [4]. The S1 subunit contains the receptor binding domain and facilitates interactions with the host cell surface receptor, Angiotensin-converting enzyme 2 (ACE2) [5,6]. The S2 subunit, activated by the host Transmembrane Serine Protease 2, harbors necessary elements for membrane fusion [7]. S protein mutations may induce conformational changes leading to increased pathogenicity [8]. We were pioneers to report the perspective role of S protein D614G (23403A>G) variant located at the S1-S2 proximal junction. The D614G mutation generates conformational changes in the protein structure which renders the FURIN cleavage site (664-RRAR-667) flexible and thus enhances viral entry [9]. Currently, the D614G mutation has become the focus of several recent studies for the use in prospective drug targeting strategies [10]. Furthermore, studies on understanding the genetic diversity and evolution of SARS-CoV-2 are emerging [11]. Admixture analyses have been conducted to understand the evolution of Beta-coronaviruses and in particular the diversification of SARS-CoV-2 [12]. Haplotype analysis have also indicated that frequencies of certain haplotypes correlate with virus pathogenicity [13].

Here we extended our study by analyzing the population genetics aspects within genome sequences of SARS-CoV-2 to understand the contiguous spread of SARS-CoV-2, its rapid evolution, and the differential severity of COVID-19 among different continents. We analyzed 2341 viral sequences deposited in GISAID including those sequenced in our lab from patients in Kuwait. We found evidence of coinfection between different viral strains in Europe and in North America but not in the other continents. We also modelled major mutations and their possible effect on the stability of the encoded protein and hence on SARS-CoV-2 virulence.

## Methods

### Retrieval of complete SARS-CoV-2 genome sequences

2431 complete SARS-CoV-2 genome sequences from infected individuals were retrieved from the GISAID database (Global Initiative on Sharing All Influenza Data) [14] (accessed on April 3^rd^, 2020) and were used for all analyses.

### Alignment and annotation of amino acid sequence variation

Multiple sequence alignment was performed using MAFFT v7.407 [15] (retree: 5, maxiter: 1000). Alignments’ gaps were trimmed using TrimAL (automated1) [16]. SeqKit [17] was used to concatenate chopped-off missing sequences using NC045512.2 as a reference. SNP-sites [18] and Annovar [19] were used to extract and annotate single nucleotide variants (SNV).

### Linkage disequilibrium and haplotype blocks analysis

PLINK2 [20] was used to extract sequence variants with minor allele frequencies (MAF) ≥ 0.5%, to estimate inter-chromosomal Linkage disequilibrium (LD), squared correlation coefficient (*r*^*2*^), and haplotype blocks. We used Haploview [21] to visualize the haplotype blocks.

To detect recombination within datasets, we tested for pairwise homoplasy index using PhiPack software [22]. Admixture 1.3.0 software [23] was used to identify genetic substructure of variants’ continental transmission as follows: variants with MAF ≥ 0.5%, variants in LD (R^2^>0.5), haplotype blocks, variants not in LD, nonsynonymous, and synonymous variants. All analyses were iterated for K=10 and cross validation errors were examined to infer an optimal K cluster. Replicate runs were further processed using CLUMPAK [24] and results for the major modes were illustrated using R software’s ggplot2 package (https://www.r-project.org/).

### Protein Structural Analysis

The crystal structures of the viral RNA-dependent RNA polymerase (RdRp, PDB ID: 6M71) [25] and the S protein (PDB ID:6VSB) [26] were used as model proteins for the structural analysis. The missing aa, invisible by Cryo-EM structure of the S-protein, were modeled-in by using SWISS-Model [27]. DynaMut web server [28] was used to predict the effect of the mutations on the proteins stability and flexibility. PyMol (Molecular Graphics System, Version 2.0, Schrodinger, LLC) was used to generate structural images.

## Results

### Detection and classification of mutations from global SARS-CoV-2 genome sequences

We analyzed 2431 high quality SARS-CoV-2 genome sequences from six continental groups. In comparison to the Wuhan reference sequence (NC045512.2), 2352 sequences showed substantial genetic differences. We identified 1010 unique missense amino acid (aa) mutations using our variant calling pipeline. 613 variants were nonsynonymous, 387 variants were synonymous, 3 variants were stop-gain, and 1 variant was a 5’-utr variant. We found only 72 variants at MAF ≥ 0.5% which were for admixture and haplotype block analysis. **Fig 1** shows the distribution of synonymous and nonsynonymous variants in each gene in SARS-CoV-2 genome with varying MAF thresholds. The genes with the highest percentage of nonsynonymous variants with MAF ≥ 0.05 are ORF3a, M, and ORF8 while the genes with the highest percentage of synonymous variants with MAF ≥ 0.05 are ORF6 and ORF10.

**Fig 1.**
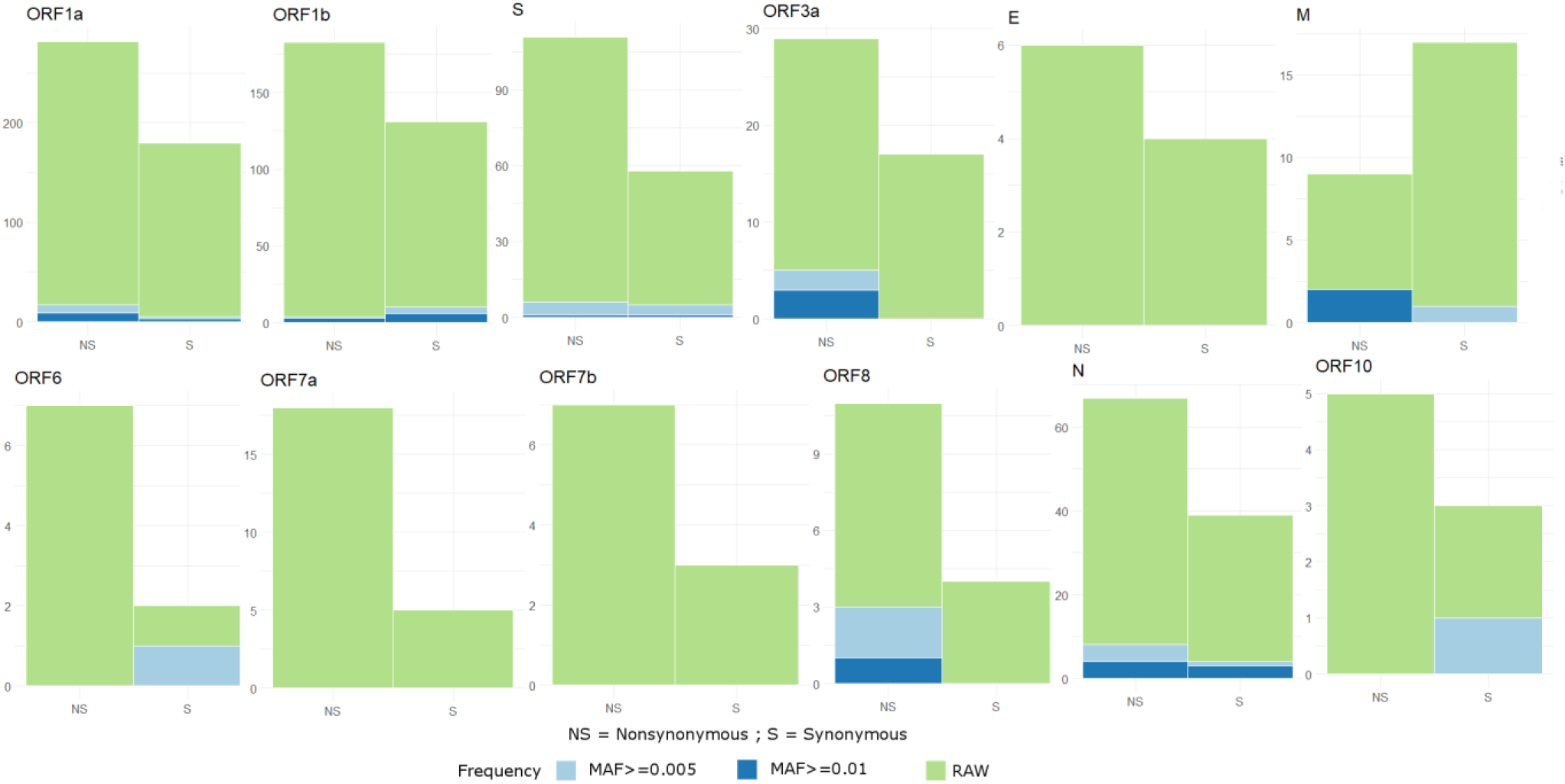
Detection and classification of mutations from GISAID SARS-CoV-2 genome sequences. Illustration of the distribution of synonymous and nonsynonymous variants for each gene in RAW, MAF ≥ 0.5%, and MAF ≥ 1% thresholds. With a threshold at MAF ≥ 0.5%, a set of 72 variants in total was observed and utilized in subsequent analysis. MAF: Minor Allele Frequency.

### Identification of SARS-CoV-2 genetic clusters in different continents

We performed Principal Component Analysis (PCA) of 2352 SARS-CoV-2 sequences as indicated in **Fig 2**. The PCA analysis gave three distinct clusters of samples based on their continent of origin. All three clusters diverge from a single point (**Fig 2**, red circle). The North American cluster showed the least viral genetic variances; unlike, samples from Asia and Oceania which harbored the most genetic diversity. Whereas the European cluster is well defined with few interspersed Asian samples which is an indication of its origin. This clustering in Europe and North America is probably associated with a founder effect where a single mutation was introduced and subsequently transmitted. This suggestion is corroborated by the fact that the collection date of the founder strain is prior to that observed in the European and the North American clusters (S1 Fig).

**Fig 2.**
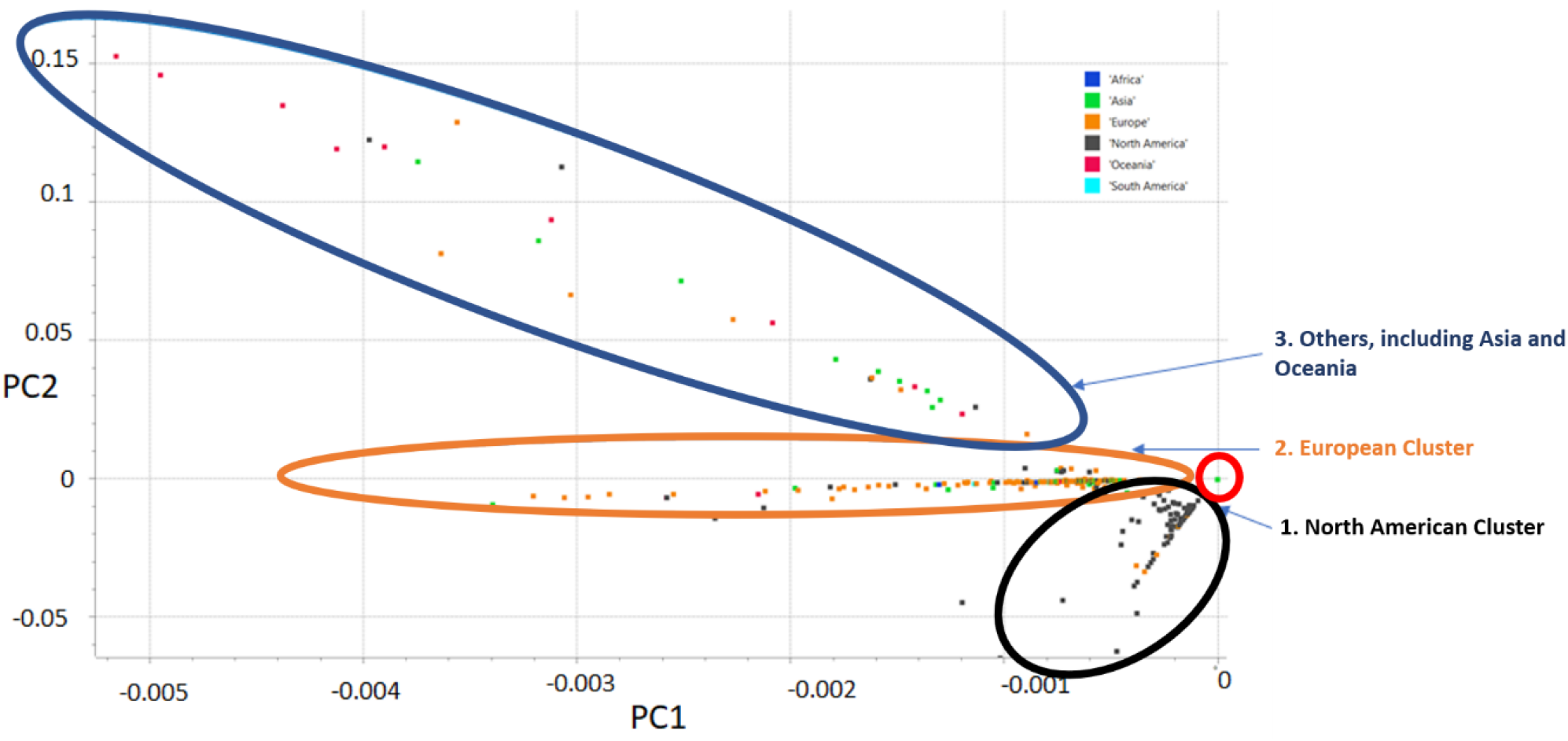
Principal Component Analysis using 2352 GISAID sequences. Principal Component Analysis of 2352 SARS-CoV-2 sequences shows three distinct clusters of color-coded samples (see the legend for their continent of origin). All three clusters diverge from a single point (red circle). The North American cluster (black oval) shows least variance among the three. The European cluster (orange oval) is well defined with few interspersed Asian samples, an indication of its origin. The third cluster (Blue oval) shows the most variance and includes samples from Oceania, Asia, and others.

Then, in order to estimate the ancestral allele frequencies and admixture proportions for each SARS-CoV-2 sample, admixture analysis was implemented. We used cross validation (CV) procedure to find best (i.e., minimal error) K for a range of iterations in RAW (comprised 1010 SNVs). S2 Fig presents the trend of CV error in RAW (A) and variants filtered for MAF≥0.5% (B). We observed a gradual reduction in CV error for iterations up to K=7 and subsequently, a pattern of an increase-decrease was observed in subsequent iterations; however, the least error was seen at K=17. Further, we verified this CV trend in a subset of variants filtered for MAF ≥0.5% (comprised of 72 SNVs). Interestingly, we again observed a gradual reduction of CV error up to K=7 (with best fit at 7) and subsequently, a trend of increasing CV error.

We created subsets of strong LD, weak LD, Haplotype block, nonsynonymous, and synonymous variants from 72 variants at K=7 across the continents. We separately performed admixture analysis on these subsets. The analysis of the detected seven datasets revealed interesting mosaic patterns (**Fig 3** A and B). Samples from Asia formed largely two clusters (C1-dominant, and C6); whereas, the samples from Europe dataset distributed into six different clusters (C1, C2-dominant, C3, C5, C6 and C7); and the North American samples formed four clusters (C1, C2, C4-dominant and C7). Similarly, African and Oceanian datasets formed two clusters each (C2, C3; with dominant C2 and C3, C5; with dominant C3 respectively) and South American dataset is mostly formed by five clusters (C1, C2, C3, C5 and C6).

**Fig 3.**
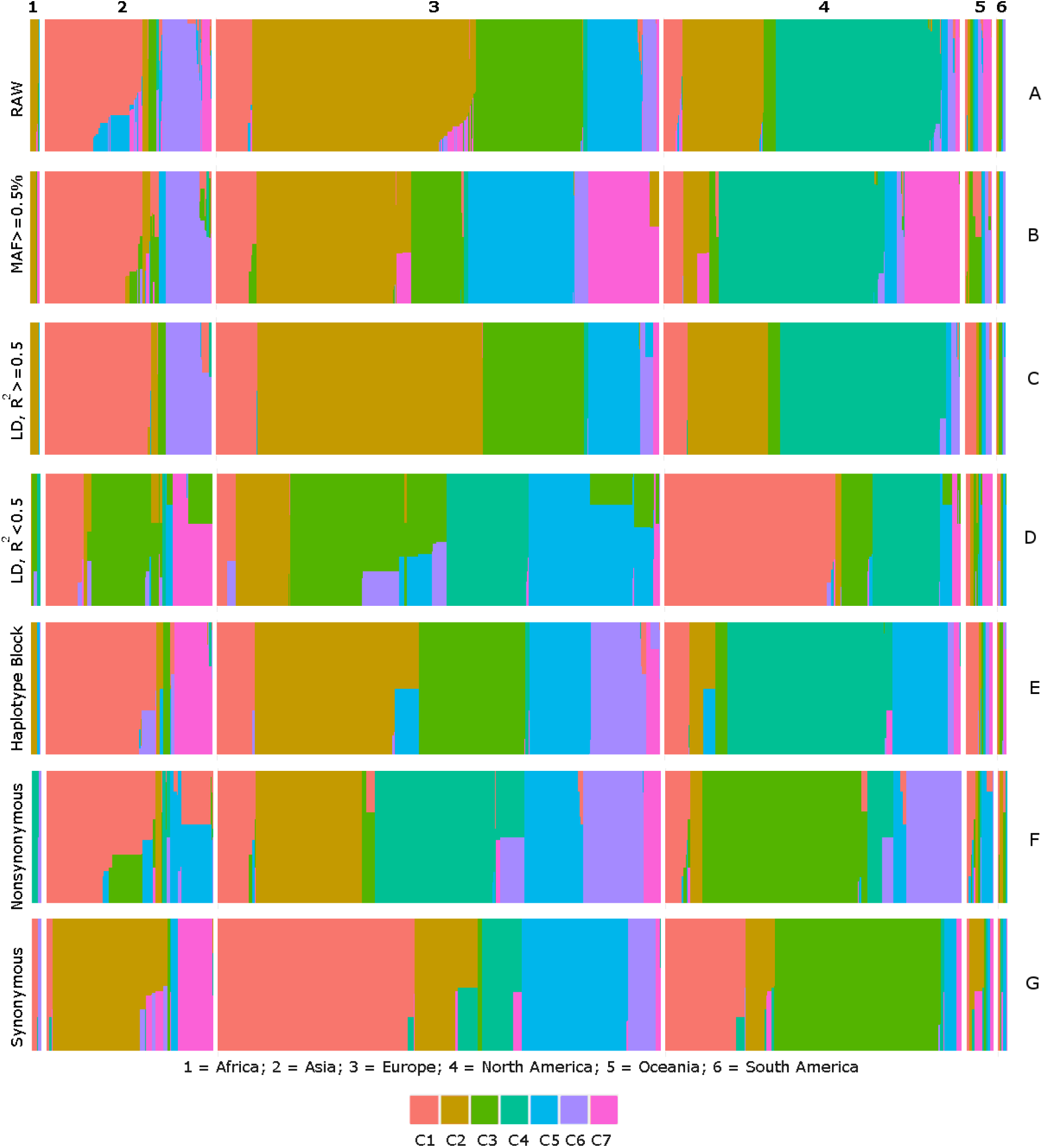
Identification of SARS-CoV-2 genetic clusters in different continents. Illustration of seven color coded (C1 to C7) genetic subdivisions of SARS-CoV-2 across continents using variants with a MAF≥0.5% frequency. Differential proportions of strong LD (C), weak LD (D), haplotype block (E), nonsynonymous (F), and synonymous (G) variants across continental datasets are shown.

In the context of LD (**Fig 3** C and D), the strong LD block is mostly observed in two clusters (C1 and C6) and the weak LD block is observed in three clusters (C1, C3, and C7) in Asia. In Europe, the strong LD block is observed in 4 clusters (C1, C2, C3, and C5) and the weak LD block is observed in five clusters (C1,C2,C3,C4, and C5; predominantly from C3 and C5). Interestingly, four clusters are common between the strong and weak LD blocks, suggesting that a makeup of four strains that dominate in Europe have significant proportion of strong and weak LD signatures in them. Likewise, in North America, both strong and weak LD are observed among three clusters (C1, C2, and C4) and (C1, C3, and C4) respectively. Further in Africa, Oceania and South America, strong LD is observed among (C2), (C1) and (C1 and C2) respectively; weak LD is observed in (C3 and C4), (C1, C2, C3, and C5), (C3 and C5) respectively. Interestingly, proportions of a Haplotype block (**Fig 3** E) identified using whole data, mostly followed pattern of strong LD of respective continents, however admixed with a weak LD strain of respective continents.

Most importantly variation in proportions of strains carrying nonsynonymous and synonymous signatures are also very evident as shown in **Fig 3** F and G. Proportions of two nonsynonymous clusters (C1 and C5; both also have admixed with each other) and two synonymous clusters are dominated in Asia. Four clusters of nonsynonymous (C2, C4, C5, and C6) and five clusters of synonymous are dominating in Europe, while in North America, three nonsynonymous (C1, C3, C6) and two synonymous (C1, C3) clusters are in higher proportions. Similarly; C4 and C6, C5, and C2 nonsynonymous clusters along with C1and C6, C2, and C5 synonymous clusters are in high proportion in Africa, Oceania, and South America respectively.

### Evidence of co-infection in the sequences from continental datasets

Here, we tested for the presence or absence of recombination among continental datasets using PhiPack software which effectively differentiates between presence or absence of recombination using three different tests “Pairwise Homoplasy Index (Phi)” (Bruen et al. 2006), “Neighbor Similarity Score (NSS)” (Jakobsen and Easteal, 1996) and “Maximum χ^2^ (MaxChi)” [29].

The results of these tests on the combined dataset suggested the possibility of co-infection at a global level. Among continental datasets, only European (NSS test, P-value = 0.001) and North American [NSS and Phi (normal), P-value = 0.007 and 0.042, respectively] sequences have shown evidence for the presence of recombination events; while African, Oceanic, South American, and Asian datasets have shown no recombination in early spread of SARS-CoV-2 (**Table 1**).

**Table 1.** Evidence of recombination in the sequences from continental datasets.

| **Dataset** | **Number of**<br>**informative variants** | **Tests to detect evidence**<br>**of recombination** | **Significance of observed**<br>**Phi statistics** |
| --- | --- | --- | --- |
| Africa (n=25) | 19 | NSS | 1 |
|  |  | MaxChi<sup>2</sup> | 0.978 |
|  |  | Phi (permutation) | 1 |
|  |  | Phi (normal) | 1 |
| Asia (n=364) | 127 | NSS | 0.113 |
|  |  | MaxChi<sup>2</sup> | 0 |
|  |  | Phi (permutation) | 0.493 |
|  |  | Phi (normal) | 0.305 |
| Europe (n=1132) | 276 | **NSS** | **0.001** |
|  |  | MaxChi<sup>2</sup> | 0.208 |
|  |  | Phi (permutation) | 0.343 |
|  |  | Phi (normal) | 0.223 |
| North America<br>(n=738) | 194 | **NSS** | **0.007** |
|  |  | MaxChi<sup>2</sup> | 0.061 |
|  |  | Phi (permutation) | 0.060 |
|  |  | **Phi (normal)** | **0.042** |
| South America (n=24) | 24 | NSS | 0.596 |
|  |  | MaxChi<sup>2</sup> | 0.680 |
|  |  | Phi (permutation) | 0.717 |
|  |  | Phi (normal) | 0.454 |
| Oceania (n=69) | 50 | NSS | 1 |
|  |  | MaxChi<sup>2</sup> | 0.502 |
|  |  | Phi (permutation) | 0.872 |
|  |  | Phi (normal) | 0.361 |
| Combined (n=2352) | 554 | **NSS** | **0.003** |
|  |  | **MaxChi<sup>2</sup>** | **0.001** |
|  |  | **Phi (permutation)** | **0.008** |
|  |  | **Phi (normal)** | **0.015** |

Results of NSS, MaxChi2, Phi (permutation) and Phi (normal) tests using pairwise homoplasy index test available from PhiPack software on the combined dataset of all the 2352 samples. Significant P-values suggest the possibility of co-infection on a global level. European (NSS test, P-value of 0.001) and North American (NSS and Phi(normal), P-value of 0.007, 0.042 respectively) show evidence for the presence of recombination events, while African, Oceanic, South American, and Asian datasets show no recombination in early spread of SARS-CoV-2 in respective continents.

### Estimation of haplotype blocks in continental samples

We carried out this analysis in two ways; first by comparing haplotype blocks obtained from combined dataset of variants with MAF ≥0.5% with each continental dataset. We did this analysis because continental datasets of Africa (n=25), Oceania (n=69), and South America (n=24) were very small to infer high confident LD blocks from respective datasets alone. Hence, we first compared LD blocks obtained from the combined dataset and then observed LD among the same variants in each continental dataset. The second way of analysis was by directly estimating haplotype blocks in each continental dataset with large sample size such as Asia, Europe, and North America.

From the first analysis, we observed that LD block, obtained from the combined dataset, varies among continental datasets. S3 Fig illustrates the extent of LD variation in haplotype block of combined dataset in each continental dataset. Examination of variant allele frequency at haplotype block suggested a clear variation in allele frequency between continental datasets. **Table 2** shows MAF of 18 variants involved in haplotype block of combined dataset in each continental dataset. These differences called for the second approach of estimating haplotype blocks and the extent of LD between variants directly from each continental dataset having large sample size. Surprisingly, we observed different sets of variants in haplotype blocks, different length of haplotype blocks, and differences in nonsynonymous composition in haplotype blocks among the three continents datasets (**Fig 4**). **Table 3** describes characteristics of the haplotype blocks observed in the datasets from Asia, Europe, and North America.

**Table 2.** MAF distribution of 18 variants involved in haplotype block of combined dataset in each continental data.

| SNV | Minor allele frequency |  |  |  |  |  |  | Functional consequence | Gene |
| --- | --- | --- | --- | --- | --- | --- | --- | --- | --- |
|  | Africa | Asia | Europe | North America | South America | Oceania | Combined |  |  |
| 241CT | 0.08 | 0.0811 | 0.2507 | 0.3231 | 0.4583 | 0.1905 | 0.49 | downstream | 5'-UTR |
| 1059CT | 0.24 | 0.019 | 0.1435 | 0.1865 | 0 | 0.0289 | 0.1313 | nonsynonymous | ORF1a |
| 3037CT | 0.08 | 0.0760 | 0.2502 | 0.313 | 0.4583 | 0.1884 | 0.4804 | synonymous | ORF1a |
| 8782CT | 0.04 | 0.2304 | 0.0424 | 0.4197 | 0.2917 | 0.1884 | 0.2484 | synonymous | ORF1a |
| 11083GT | 0.04 | 0.269 | 0.1339 | 0.0531 | 0.125 | 0.4928 | 0.1416 | nonsynonymous | ORF1a |
| 14408CT | 0.08 | 0.0760 | 0.25 | 0.3148 | 0.4583 | 0.1884 | 0.481 | nonsynonymous | ORF1b |
| 14805CT | 0.08 | 0.0047 | 0.1366 | 0.0224 | 0.3333 | 0.1159 | 0.0787 | synonymous | ORF1b |
| 17747CT | 0 | 0 | 0.0106 | 0.4616 | 0 | 0.0579 | 0.174 | nonsynonymous | ORF1b |
| 17858AG | 0 | 0 | 0.0097 | 0.4418 | 0 | 0.0579 | 0.1798 | nonsynonymous | ORF1b |
| 18060CT | 0 | 0.0166 | 0.0088 | 0.4382 | 0 | 0.0579 | 0.1829 | synonymous | ORF1b |
| 23403AG | 0.08 | 0.0783 | 0.25 | 0.3121 | 0.4583 | 0.1884 | 0.4808 | nonsynonymous | S |
| 25563GT | 0.32 | 0.0213 | 0.1851 | 0.2262 | 0 | 0.0434 | 0.1647 | nonsynonymous | ORF3a |
| 26144GT | 0.04 | 0.1119 | 0.1293 | 0.0211 | 0.125 | 0.1594 | 0.0923 | nonsynonymous | ORF3a |
| 27046CT | 0 | 0 | 0.1114 | 0.0013 | 0.125 | 0.0144 | 0.0538 | nonsynonymous | M |
| 28144TC | 0.04 | 0.2304 | 0.0407 | 0.4185 | 0.2917 | 0.1884 | 0.2483 | nonsynonymous | N |
| 28881GA | 0 | 0.0381 | 0.2396 | 0.0423 | 0.375 | 0.0869 | 0.1378 | nonsynonymous | N |
| 28882GA | 0 | 0.0381 | 0.2396 | 0.0410 | 0.375 | 0.0869 | 0.1374 | synonymous | N |
| 28883GC | 0 | 0.0381 | 0.2396 | 0.0410 | 0.375 | 0.0869 | 0.1374 | nonsynonymous | N |

Display of minor allele frequency for each variant in different continents, the functional consequence of these variants, and their corresponding genes. (SNV- single nucleotide variant)

**Table 3.** Characteristics of haplotype blocks estimated from three continental datasets.

| Dataset | Haplotype<br>block<br>start | Haplotype<br>block end | Length<br>(in kb) | Number<br>of<br>variants | Number of<br>nonsynonymous<br>variants | Variant | MAF |
| --- | --- | --- | --- | --- | --- | --- | --- |
| Asia | 3037 | 23403 | 20.367 | 5 | 3 | 3037CT | 0.076 |
|  |  |  |  |  |  | 8782CT | 0.23 |
|  |  |  |  |  |  | **11083GT** | **0.269** |
|  |  |  |  |  |  | **14408CT** | **0.076** |
|  |  |  |  |  |  | **23403AG** | **0.078** |
| Europe | 241 | 28883 | 28.643 | 17 | 10 | 241CT | 0.25 |
|  |  |  |  |  |  | **1059CT** | **0.143** |
|  |  |  |  |  |  | **1440GA** | **0.052** |
|  |  |  |  |  |  | 3037CT | 0.25 |
|  |  |  |  |  |  | **11083GT** | **0.134** |
|  |  |  |  |  |  | **14408CT** | **0.25** |
|  |  |  |  |  |  | 14805CT | 0.136 |
|  |  |  |  |  |  | 15324CT | 0.062 |
|  |  |  |  |  |  | 17247TC | 0.0689 |
|  |  |  |  |  |  | 20268AG | 0.0734 |
|  |  |  |  |  |  | **23403AG** | **0.25** |
|  |  |  |  |  |  | **25563GT** | **0.185** |
|  |  |  |  |  |  | **26144GT** | **0.129** |
|  |  |  |  |  |  | **27046CT** | **0.111** |
|  |  |  |  |  |  | **28881GA** | **0.239** |
|  |  |  |  |  |  | 28882GA | 0.239 |
|  |  |  |  |  |  | **28883GC** | **0.239** |
| North<br>America | 241 | 8782 | 8.54 | 4 | 1 | 241CT | 0.323 |
|  |  |  |  |  |  | **1059CT** | **0.186** |
|  |  |  |  |  |  | 3037CT | 0.313 |
|  |  |  |  |  |  | 8782CT | 0.419 |
|  | 14408 | 28144 | 13.737 | 7 | 6 | **14408CT** | **0.3148** |
|  |  |  |  |  |  | **17747CT** | **0.462** |
|  |  |  |  |  |  | **17858AG** | **0.442** |
|  |  |  |  |  |  | 18060CT | 0.438 |
|  |  |  |  |  |  | **23403AG** | **0.312** |
|  |  |  |  |  |  | **25563GT** | **0.226** |
|  |  |  |  |  |  | **28144TC** | **0.418** |

Characteristics presentation of haplotype blocks estimated from the datasets obtained from Asia, Europe and North America. Nonsynonymous variants are shown with bold font.

**Fig 4.**
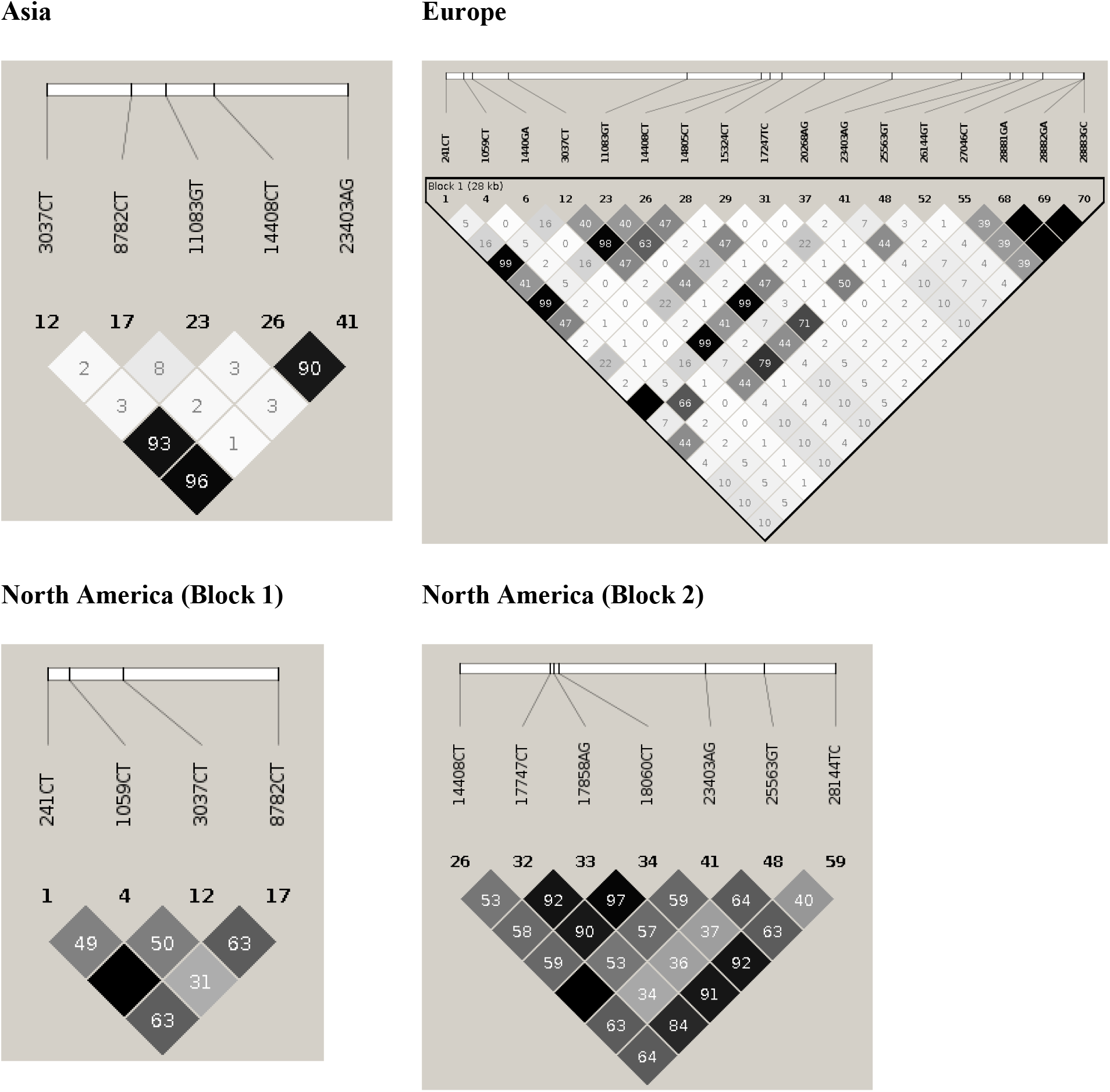
Estimation of haplotype blocks in continental samples. Haplotype block estimation and extent of Linkage disequilibrium observed between variants in Asia, Europe, and North America, identified a single block with different lengths in Asia and Europe, while in North America two blocks were identified.

### Structural analysis of SARS-CoV-2 mutations

#### D614G mutation

Wrapp et al. recently solved the cryo-EM structure of the S protein with 3.5 Å resolution [26] (**Fig 5** A). The S protein has many flexible loop regions that were not visible in the structure, including the RRAR cleavage site. Therefore, we modeled-in the cleavage site and the undetected flexible regions using the cryo-EM structure PDB ID: 6M71 as a scaffold [27]. **Fig 5** B shows the overlay of the S protein from PDB ID:6VSB with the modelled S protein, with an RMSD of 0.25 Å. As shown in the overlay figure, there are more loops present in the modelled structure that were not detected by Cryo-EM. The RRAR cleavage site (**Fig 5** C) shows a high surface accessible region, where the viral protein can attach to the host protein. As such, any mutations on the S protein especially close to RRAR site might alter its activity. D614G is a mutation believed to increase SARS-CoV-2 virulence [9]. One possibility is that the change from a negatively charged aspartate to a non-polar glycine may modify the structure and therefore the function of the protein. Charged amino acids form ionic and hydrogen bonds (H-bond) through their side chains and stabilize proteins [30]. The targeted aspartate is present in the loop region, therefore a mutation to a glycine would cause unfolding of the loop and possibly render it more flexible making the FURIN cleavage site more accessible.

**Fig 5.**
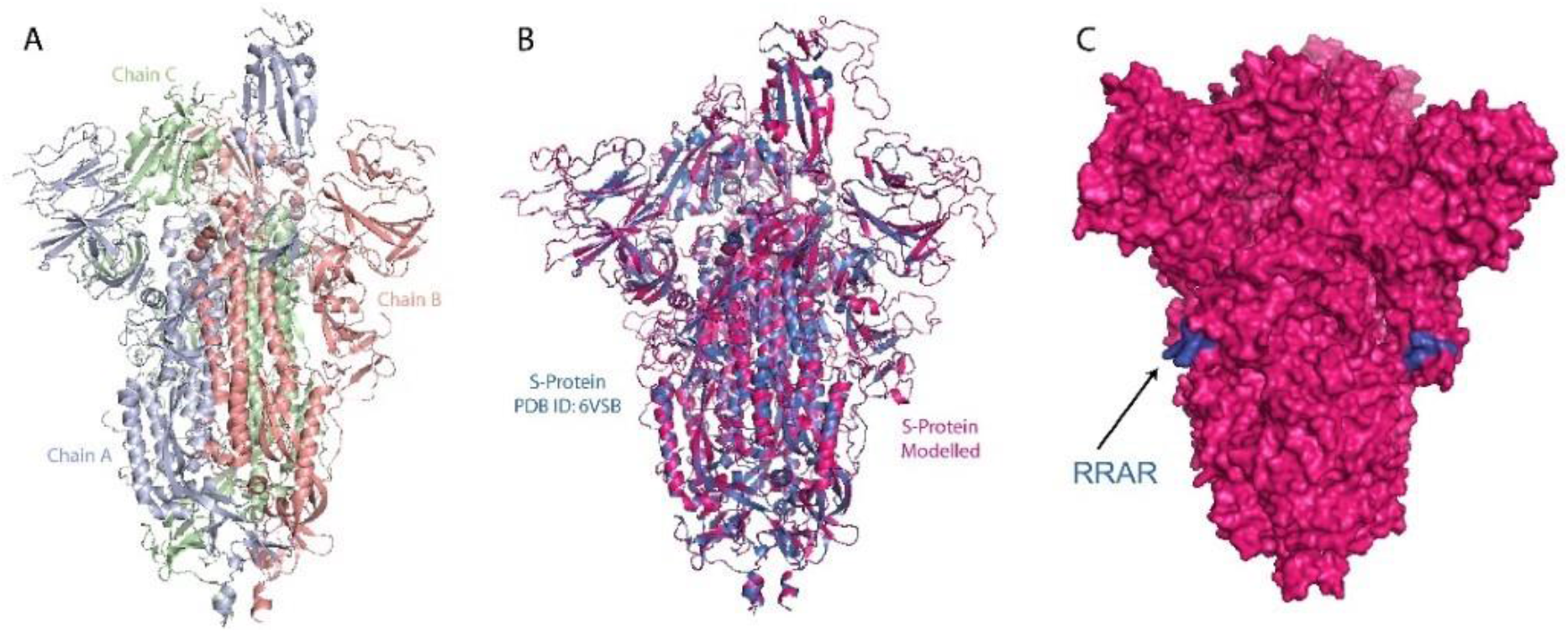
3D modeling of SARS-CoV-2 Spike protein. (A) Trimeric structure of SARS-CoV-S spike like protein (PBD:6VSB). (B) Overlay of the SARS-CoV-S spike like protein (PBD ID: 6VSB, blue) with the modelled SARS-CoV-2 S protein (PDB ID: 6M71, magenta). (C) The surface of the modelled S-protein with the RRAR FURIN cleavage site (blue)

D614 is in close vicinity to T859 of the adjacent monomer’s S2 (Chain B) where they can form a H-bond (**Fig 6**) through both sidechains. In addition, backbone H-bonds can be formed with A646 of the same chain. It was documented that S2 domains alter their structure after FURIN site cleavage [31]. Therefore, the mutation of D614 to G might weaken the stability of S2 and make cell entry more aggressive. It is probably the loss of the H-bond between D614 (S1/Chain A) and T859 (S2/Chain B) that stops the hinging of the S2 domain making it more flexible in the transition state when interacting with the host cell receptor. Another possibility would be that the mutation to G and the loss of the H-bond to the adjacent chain made the protein more flexible. A thermodynamic analysis showed that D614G mutation resulted in slightly destabilizing the protein with a ΔΔG: −0.086 kcal/mol and in increasing in vibrational entropy ΔΔSVib 0.137 kcal.mol^-1^.K^-1^ as seen in **Fig 7** A where the red parts indicate more flexibility. Since this mutation will occur on the trimeric structure of the S-protein, all the three domains will be more flexible. Such flexibility will render the FURIN cleavage site more accessible which is concomitant with the virulence of the D614G mutation. Furthermore, the flexibility observed in the ribosomal binding domain region (**Fig 7**) may facilitate the binding of ACE2 to the S protein [26].

**Fig 6.**
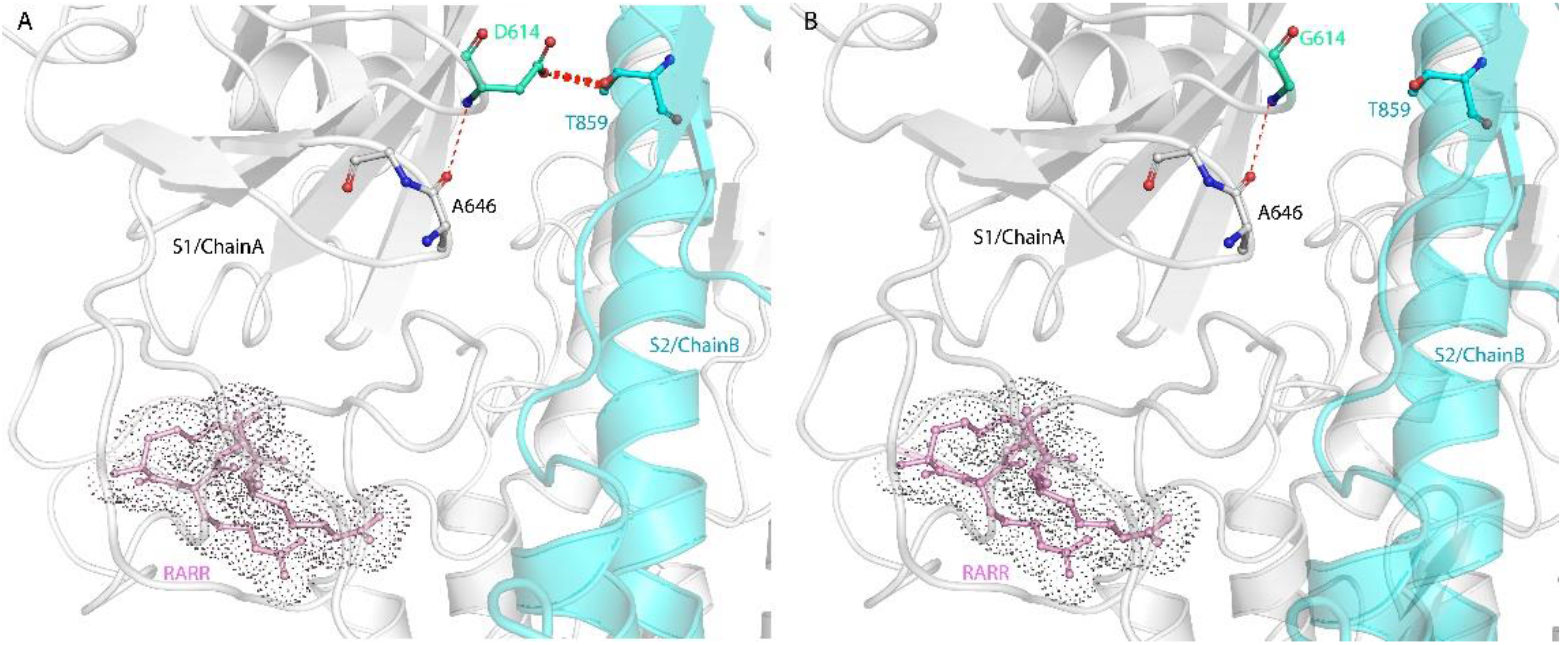
3D modeling of SARS-CoV-2 Spike protein showing suggested bonds for D614. (A) Suggested hydrogen bond (red dashed lines) of D614 (S1 domain chain A) with T859 (S2 domain chain B) and D614 and A646 of S1 domain chain A. (B) The suggested hydrogen bond can be disrupted with the D614G mutation altering the activity of the protein.

**Fig 7.**
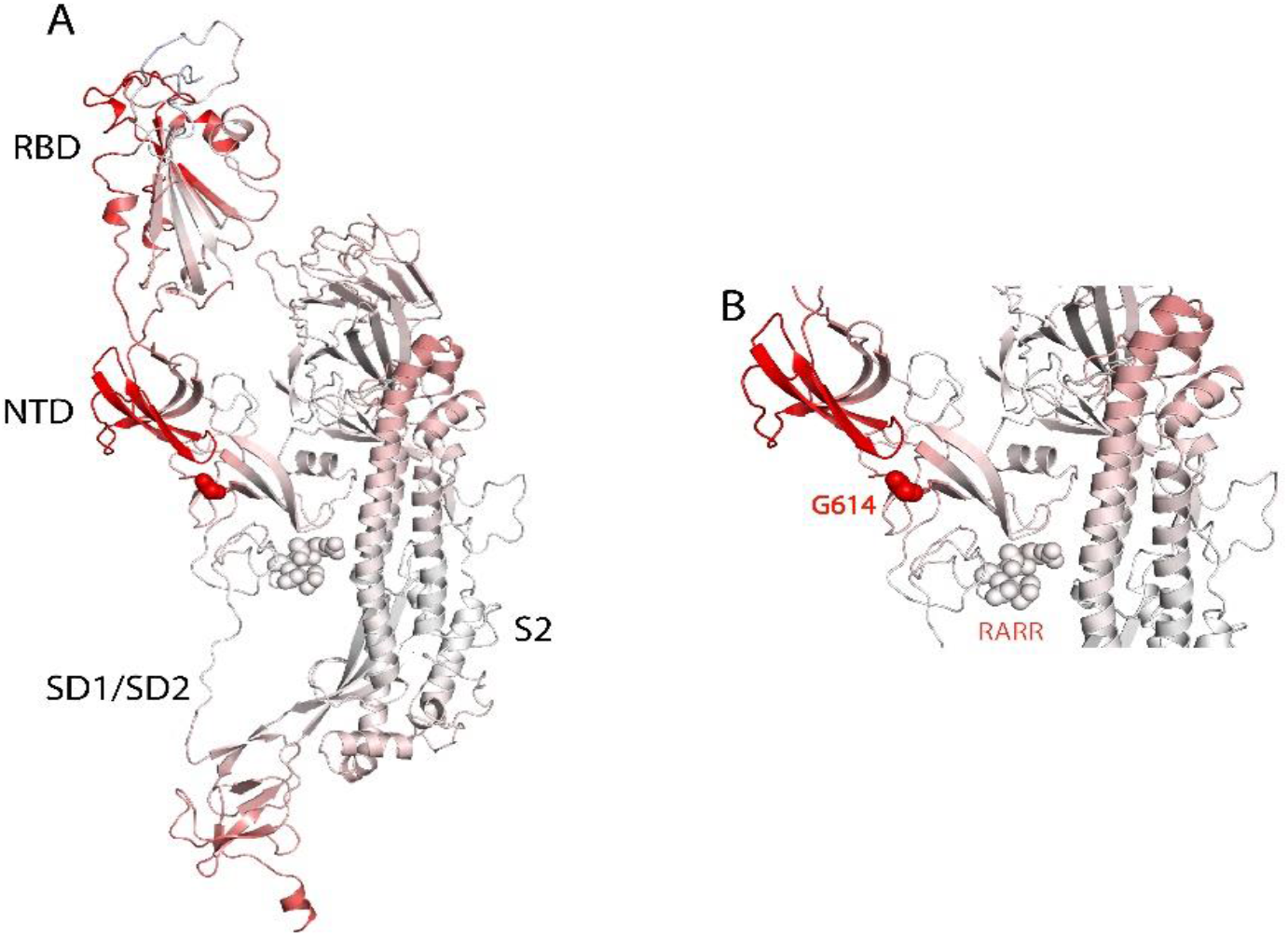
3D modeling of G614 mutation. (A) S-protein monomer 6VSB D614G, the red region of the protein depicts the more flexible region of the protein due to the D614G mutation with a decreased stability of ΔΔG: −0.086 kcal/mol and in increase in vibrational entropy ΔΔSVib 0.137 kcal.mol-1.K-1. (B) The G614 mutation (in red) suggests an increased flexibility of the region of the S-protein.

#### P314L mutation

ORF1a and ORF1b produce a set of non-structural proteins (nsp) which assemble to facilitate viral replication and transcription (nsp7, nsp8 and nsp12) [32]. The nucleoside triphosphate (NTP) entry site and the nascent RNA strand exit paths has positively charged aa, is solvent accessible, and is conserved in SARS-CoV-2 (**Fig 8**). P314L mutation is positioned on the interface domain of the RdRp (or nsp12) between A250-R365 residues. Previous studies have shown that the interface domain has functional significance in the RdRp of *Flavivirus*. In addition, when polar or charged residue mutations were introduced into these sites, viral replication levels were significantly affected [33]. Thus, mutations on nsp12 interface residues may affect the polymerase activity and RNA replication of SARS-CoV-2. Proline is often found in very tight turns in protein structures and can also function to introduce kinks into α-helices. In **Fig 9**, we investigated the proposed intermolecular bonds that P314 can make, where the backbone COO^-^ group of proline can form H-bonds with the backbone NH groups of T and S or the OH^-^group of S side chain. Whereas the pyrrolidine forms hydrophobic interactions with the W268 and F275. The mutation to leucine tightens the structure and reduces the flexibility with an increase in ΔΔG: 0.717 kcal/mol and a decrease in vibrational entropy to ΔΔSVib ENCoM: −0.301 kcal.mol^-1^.K^-1^ (**Fig 10**). Furthermore, leucine possesses a non-polar side chain, seldomly involving catalysis, which can play a role in substrate recognition such as binding/recognition of hydrophobic ligands. L314 backbone COO-forms a H-bond with the sidechain OH-group of S325, in addition to a hydrophobic interaction with W68 and L270. L270 is positioned on top of the flexible loop region, therefore forming a hydrophobic interaction, displacing any possibility of water molecules entering the looped region thus making it more compact. The overall improved stability of RdRp can make it more efficient in RNA replication and hence increasing SARS-CoV-2 virulence.

**Fig 8.**
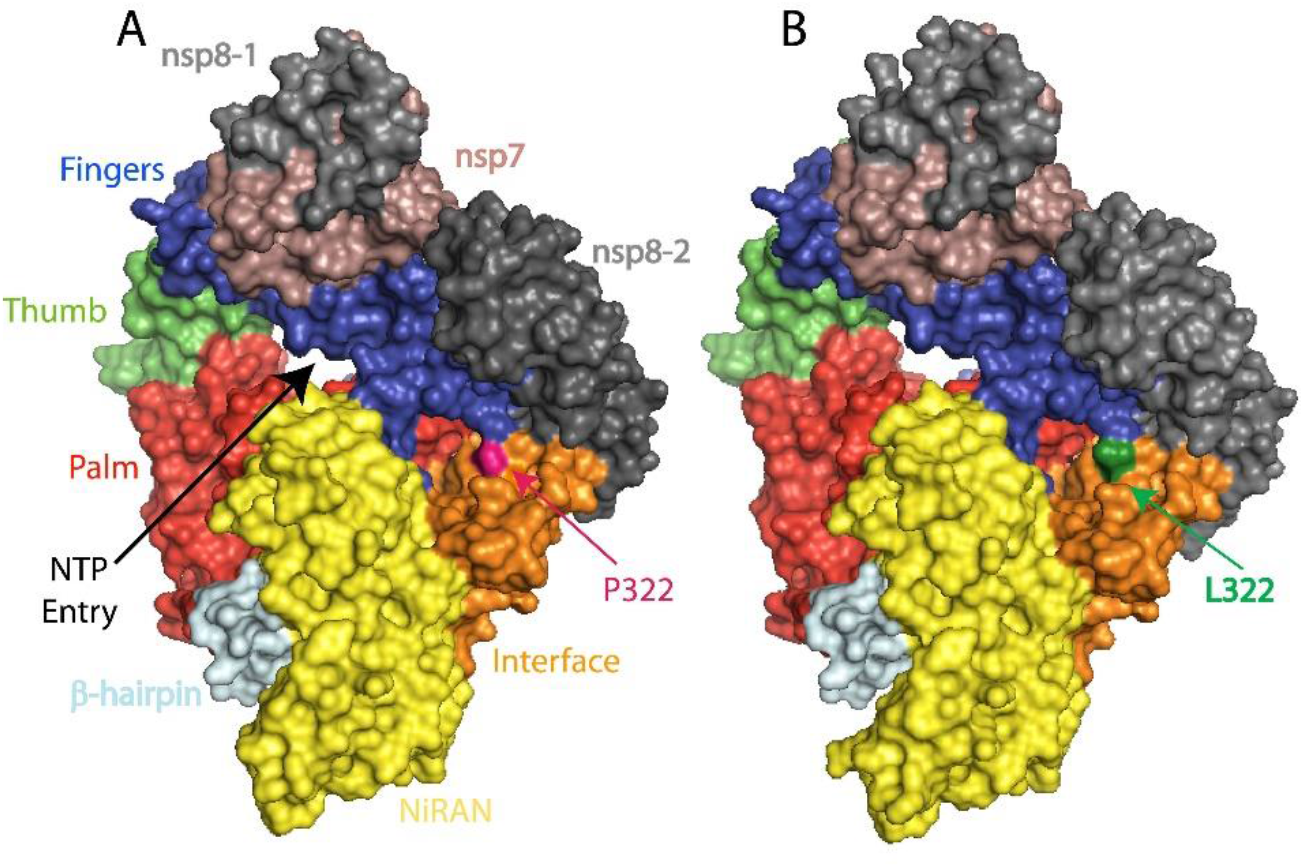
SARS-CoV-2 RNA-dependent RNA polymerase structure in complex with nsp7 and two nsp8. Viral RNA template entry and the NTP entries are shown in black arrow heads, prospective route for the release of RNA template and product after replication is shown in black arrow and two dash black arrows. The active site is a large groove with several structural pockets. (A) Wild type RdRp complex P314 (in pink) (B) L314 mutation (in green color). RdRp-RNA-dependent RNA polymerase.

**Fig 9.**
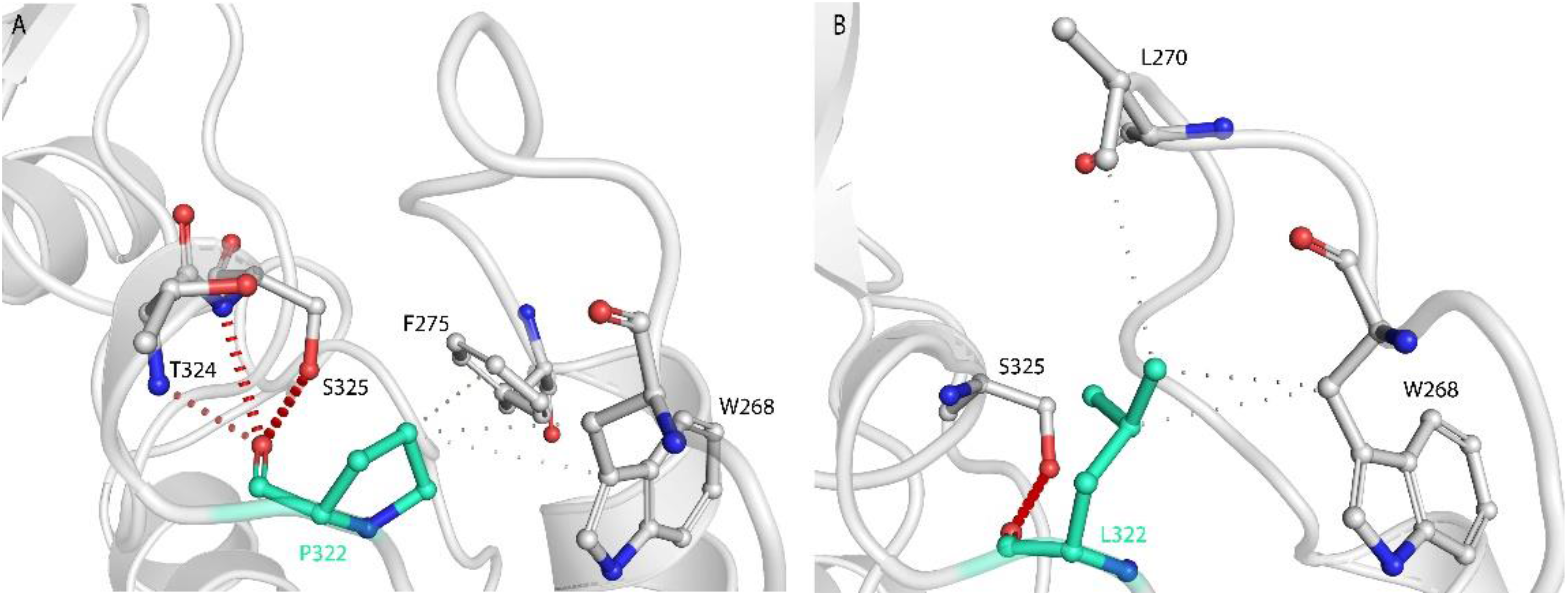
3D modeling of P314L mutation. (A) Suggested bonding network of P314 where the COO-group might form H-bonds with the backbone NH of T324 and S325 and the side chain of S325. The white dashed lined depict the hydrophobic interactions between P314 and W268 and F275. (B) L314 forms a H-bond with the side chain of S325 and forms a hydrophobic interaction with L270 which is at the curve of the loop bringing making that region more compact.

**Fig 10.**
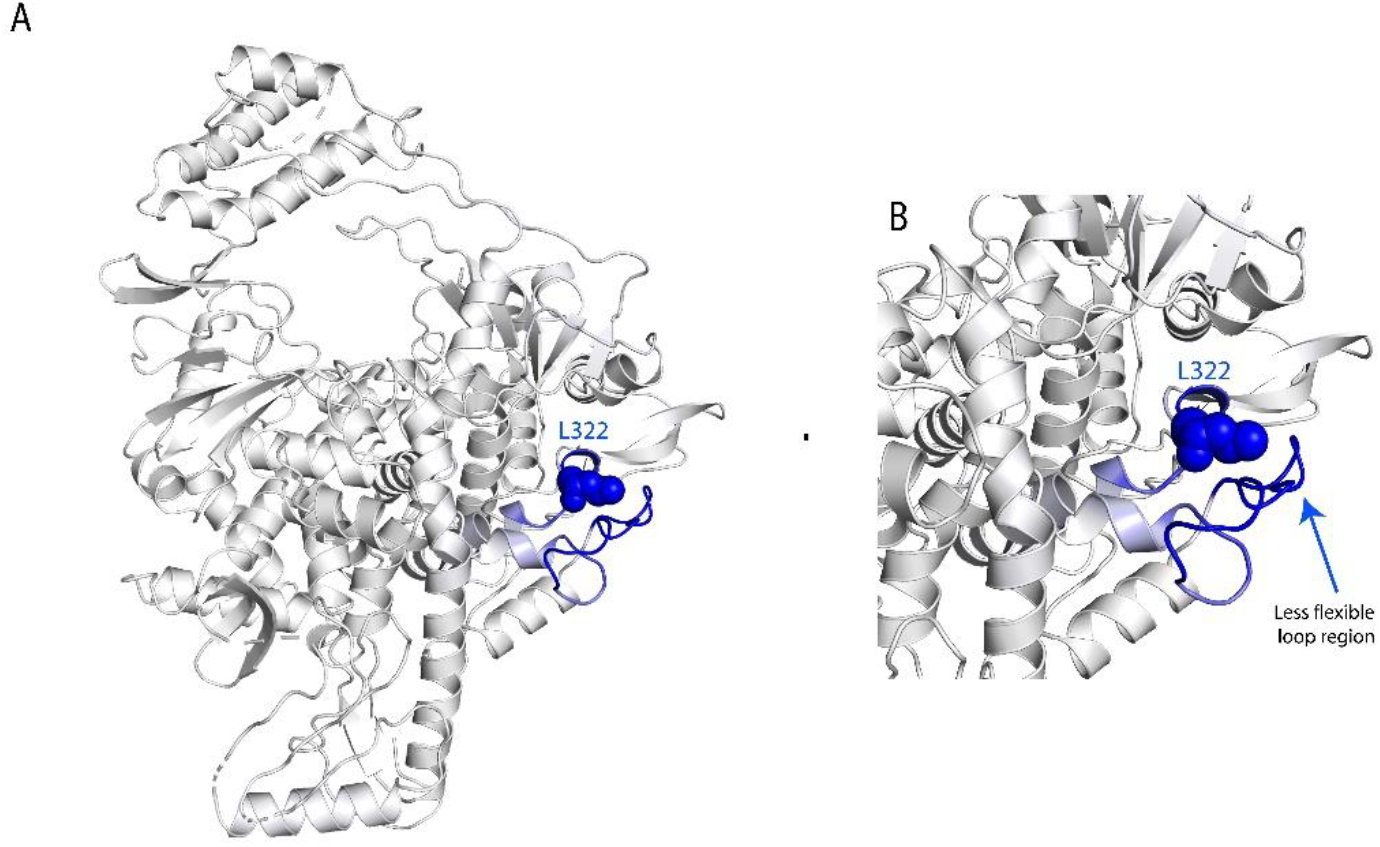
3D depiction of the less relaxed loop caused by L314 mutation. (A) RNA-dependent RNA polymerase structure, the blue region of the protein depicts the more rigid region due to the mutation P314L, with an increase in stability of in ΔΔG: 0.717 kcal/mol and a decrease in vibrational entropy to ΔΔSVib ENCoM: −0.301 kcal.mol-1.K-1. (B) The less flexible loop region because of the tight hydrophobic interaction between L314 (L322 in structure) and hydrophobic residues in the moiety.

## Discussion

Our pairwise homoplasy index tests suggest that, among continental datasets, European and North American sequences have shown evidence for the presence of recombination events (P-value = 0.001 and 0.007 respectively); while African, Oceanic, South American, and Asian datasets have shown no recombination events till now. This again shows that the European and North American continents are at higher risk of having super evolved viruses that can co-infect their hosts. Thus far, these recombination effects might also lead to the deletion of big portions of RNA such as the one reported in a recent article, where an 81-nucleotide deletion was detected in SARS-CoV-2 ORF7a [34]. Similar deletions might give rise to attenuated viruses and aid vaccine design.

Depending on variance seen within the clustered samples, PCA analysis indicated that clustering in Europe and North America is probably associated with a founder effect where a single mutation was introduced and subsequently transmitted; 23403AG in Europe and 28144TC in North America. Admixture analysis identified differing number of clusters of viral strains in different continents: Europe with six clusters, South America with five, North America with four, and Africa and Asia with two each. Both strong and weak LD blocks are seen among clusters of strains in every continent; the four dominant strains in Europe have significant proportion of strong and weak LD signatures in them. Proportions of strains carrying missense variants over nonsynonymous variants differ among continents – five clusters of missense variants are dominant in Europe, three in North American while two clusters of missense variants are dominant in Asia, Africa, and South America. In regard to recombination patterns, European and North American continents showed evidence for the presence of recombination events among SARS-CoV-2 genomes which give indication of continuing evolution among SARS-CoV-2 viral strains.

Admixture analysis has shown 7 different strains with differential segregation of alleles in SARS-CoV-2 isolates. Upon constricting variants with strong LD, the proportional assignment did not change in Africa and Asia, whereas it changed in Europe, North America, Oceania, and South America. In fact, proportions in Europe (C2 & C3) and North America (C2 & C4) increased excessively suggesting that strong LD sites are present in more than one strain in each of these two continents. Presence of an un-admixed block pattern of strong LD between strains suggests that LD sites are not broken by significant recombination seen in these two continents. This is either due to their physical distance or natural selection. On the contrary, weak LD sites have shown clear admixture between strains. Strikingly, each continent is dominated by different set of nonsynonymous clusters such as Africa by C4, Asia by C1, Europe by C2, North America by C3, Oceania by C5, and South America by C2. This is also evident from the allele frequency variation seen in each continent.

Further, continental-wise haplotype block estimation enabled us to identify variation of linked nonsynonymous and synonymous sites in Asia, Europe, and North America. Although selection primarily acts on variation that undergo amino acid change, many synonymous variants were observed in haplotype blocks. This suggests that these synonymous sites hitch-hiked along with nonsynonymous variants due to their physical proximity. Another observed interesting feature was the variation in the number of nonsynonymous sites between Asia, Europe, and North America. Asian haplotype block carried three nonsynonymous variants, Europe haplotype block carried ten nonsynonymous variants, and North American haplotype block carried seven nonsynonymous variants. This suggests that the initial strain which originated and travelled from Asia had less functional sites whereas coinfection-led recombination in Europe and North America enriched functional sites in strains.

Our preliminary structural analysis of the European strain main mutations, D614G located in the spike gene and P314L located in the RdRp gene, showed that the first mutation will render the FURIN cleavage site more accessible while the latter would increase protein stability. 73% of the European samples have both mutations segregating together; while in Africa only 11% of the sequenced viral samples have them. This is probably the reason behind the elevated mortality rates in Europe. This points out to the fact that the virus has evolved at an alarming rate by introducing two mutations that increase its chances of survival. The European strain harbors additional mutations, notably the hotspot mutations R203K and G204R that cluster in a serine-rich linker region at the RdRp. It was suggested that these mutations might potentially enhance RNA binding and replication and may alter the response to serine phosphorylation events [35], which might further exacerbates SARS-CoV-2 virulence.

We understand that other confounding factors like SARS-CoV-2 testing, socioeconomical status, the availability of proper medical services, and the burden of other diseases are important contributors to the disparities seen in mortality rates around the world. It is imperative that more comprehensive studies should be conducted as more patient data emerges from different parts of the world. We also realize that our analysis focuses only on the early spread samples and that we have to keep analyzing the viral sequences to determine if recombination is occurring between other strains and identify any new mutations specially since the world is currently fighting the second wave of the virus. Our data highlight the urgent need to correlate patients’ medical/infection history to the viral variants in order to predict in a more accurate and personalized way how different viral strains are influencing this pandemic.

## Data Availability

The datasets generated for this study are available on request to the corresponding author.

## Acknowledgements

We gratefully acknowledge the authors from originating and submitting laboratories of the sequences in GISAID’s EpiFlu™ Database on which this research is based. All data submitters may be contacted directly via www.gisaid.org.

## Supporting Information

**Supplementary Figure S1. Principal Component Analysis for the 2352 SARS-CoV-2 sequences distributed by their collection month**. Principal Component Analysis (PCA) blot based on the collection month (January, February, March) for the 2352 SARS-CoV-2 sequences extracted from GISAID data; PC1 (EV=9.7030), PC2 (EV=8.84814). Red circle indicates the founder strain that was sequenced in January.

**Supplementary Figure S2. Trend of CV error in RAW and in variants filtered for MAF ≥ 0**.**5%**. The CV for the MAF>0.5% dataset comprised 72 variants shown. K = 7 is the best fit (upon observing consistency in CV error between raw and MAF≥0.5% at K=7, optimum number of clusters 7 was selected; the inconsistency observed in RAW from K=8 may be resulting from MAF<0.5% variants), suggesting that 7 different SARS-CoV-2 strains existed in early transmission of SARS-CoV-2 across continents. (CV-cross validation procedure).

**Supplementary Figure S3. Linkage disequilibrium (LD) variation in haplotype block of combined dataset in each continental dataset**. Extent of LD variation observed in each continental dataset when haplotype block comprising the set of 18 variants identified in combined dataset were mapped to continental datasets.

