## Supplementary Material for "SARS-CoV-2: Proof of recombination between strains and emergence of possibly more virulent ones"

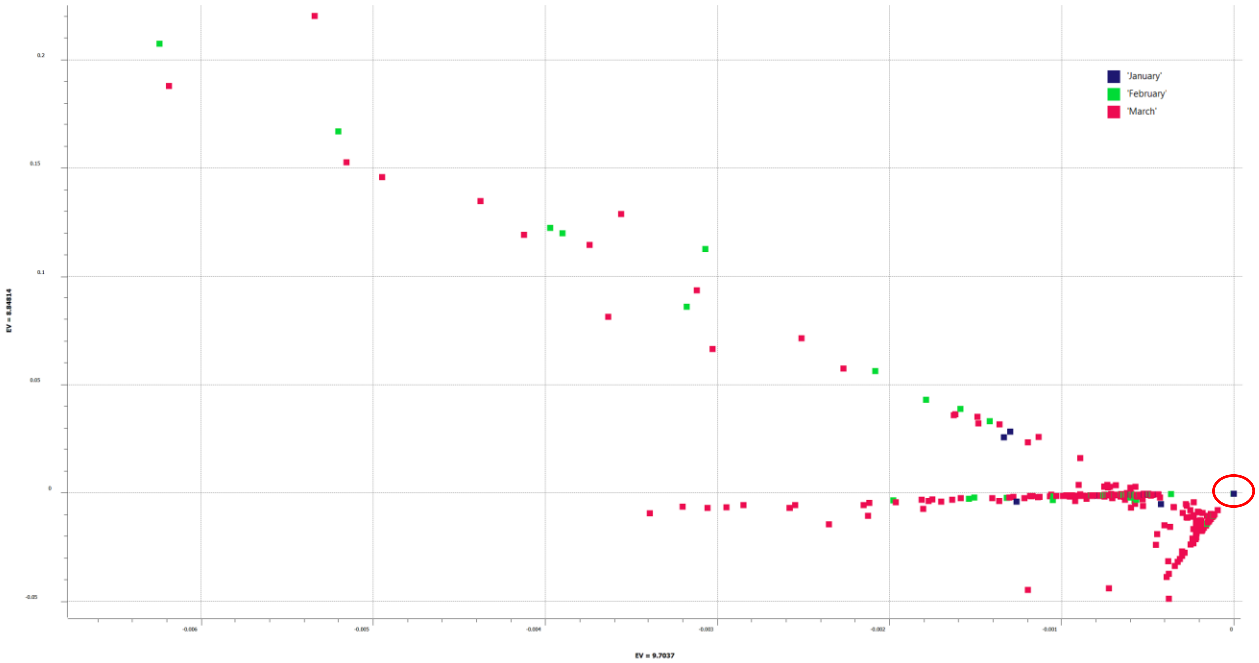


**Supplementary Figure S1. Principal Component Analysis for the 2352 SARS-CoV-2 sequences distributed by their collection month**. Principal Component Analysis (PCA) blot based on the collection month (January, February, March) for the 2352 SARS-CoV-2 sequences extracted from GISAID data; PC1 (EV=9.7030), PC2 (EV=8.84814). Red circle indicates the founder strain that was sequenced in January.

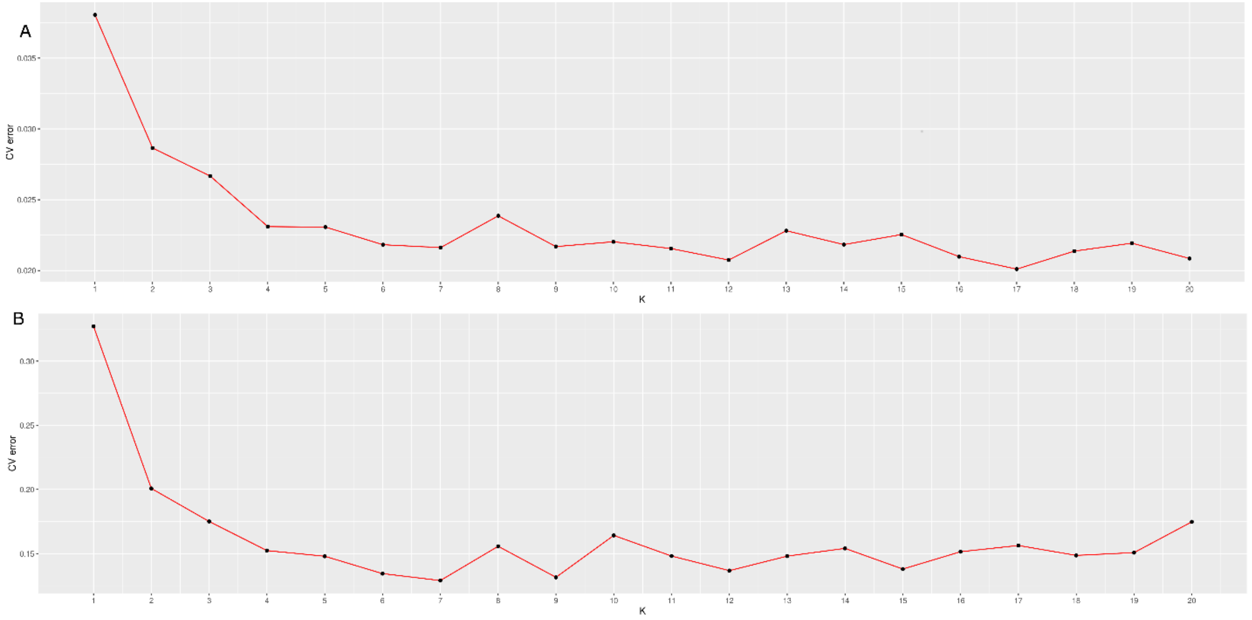


**Supplementary Figure S2. Trend of CV error in RAW and in variants filtered for MAF ≥ 0.5%.** The CV for the MAF>0.5% dataset comprised 72 variants shown. K = 7 is the best fit (upon observing consistency in CV error between raw and MAF≥0.5% at K=7, optimum number of clusters 7 was selected; the inconsistency observed in RAW from K=8 may be resulting from MAF<0.5% variants), suggesting that 7 different SARS-CoV-2 strains existed in early transmission of SARS-CoV-2 across continents. (CV-cross validation procedure)

### LD block from combined dataset with MAF≥0.5% set

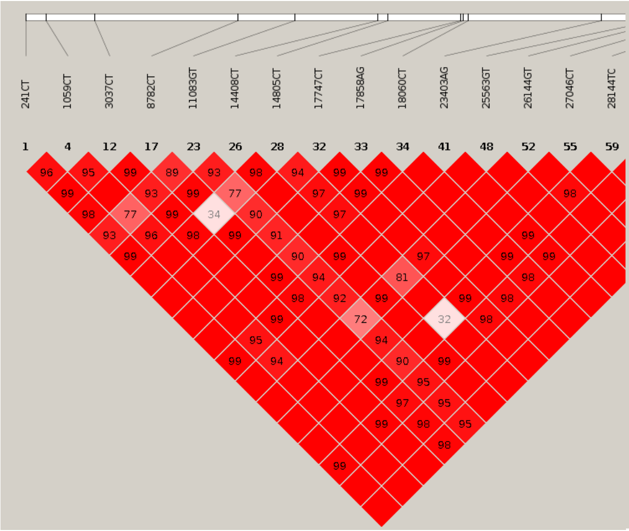


### North America

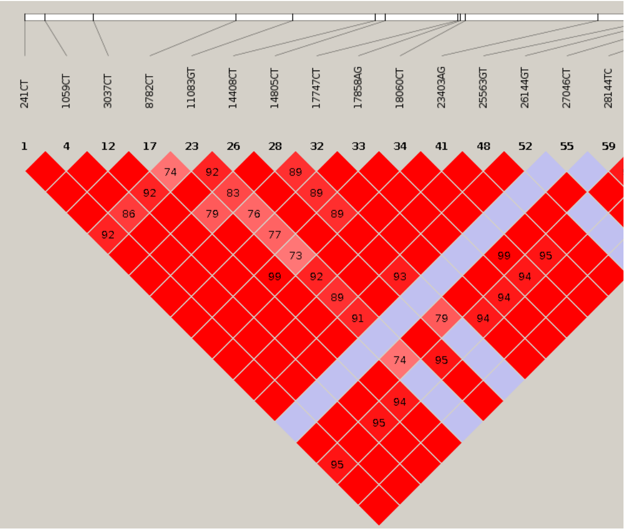


### South America

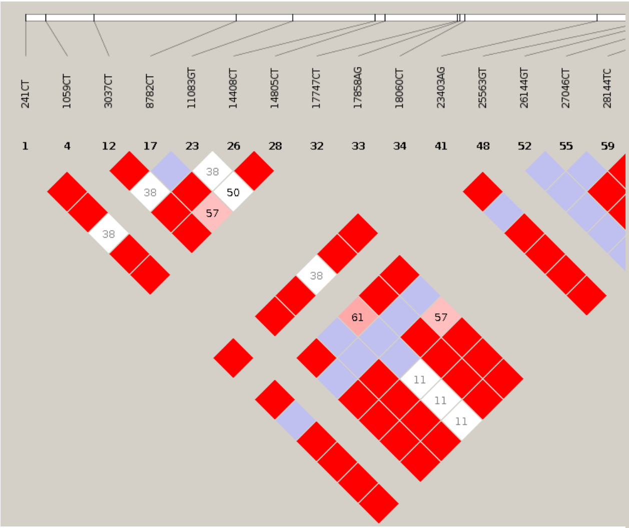


### Oceania

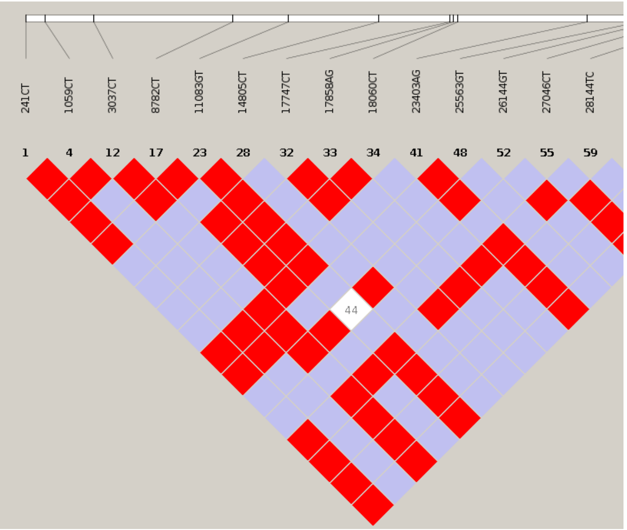

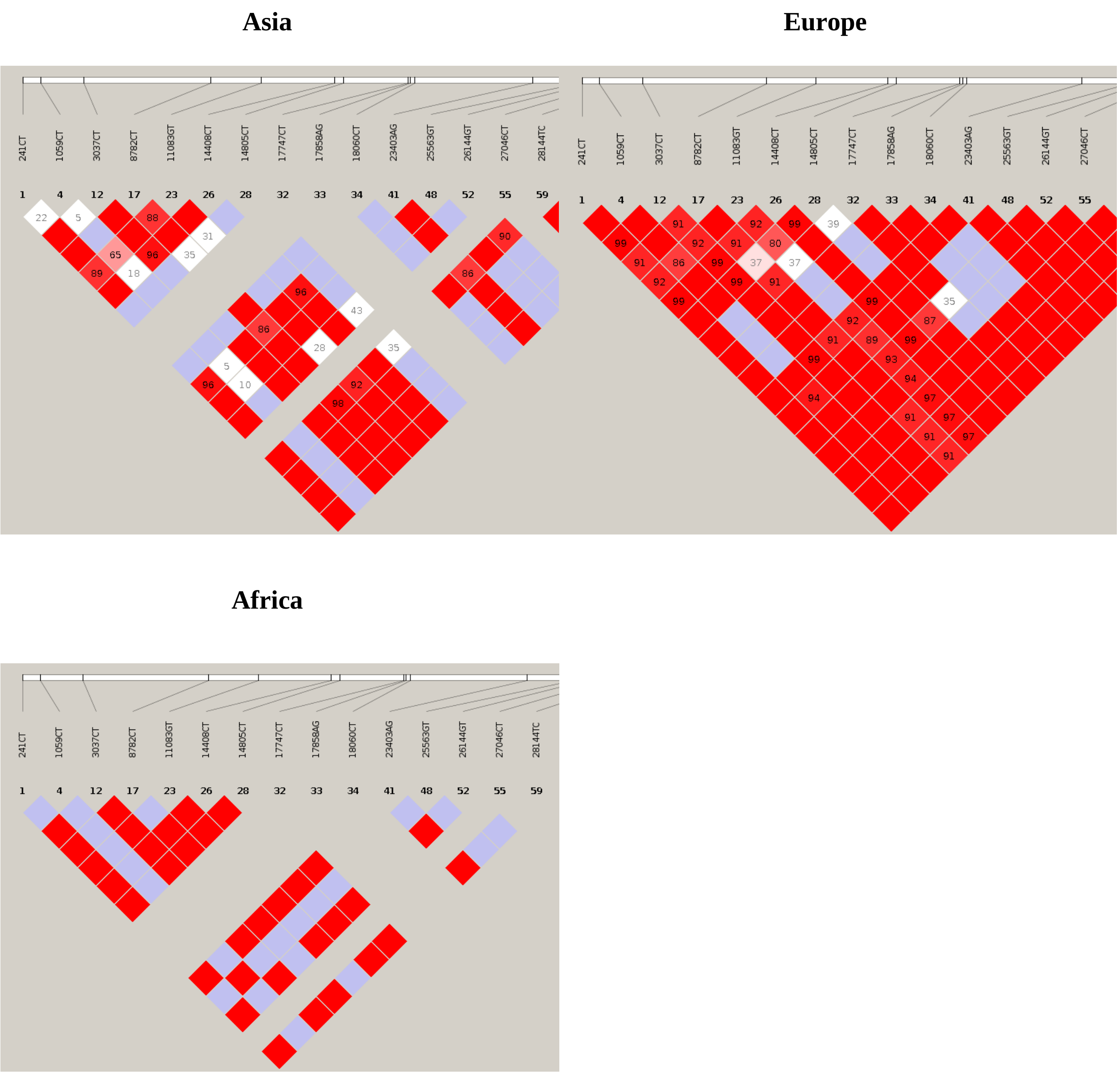


**Supplementary Figure S3. Linkage disequilibrium (LD) variation in haplotype block of combined dataset in each continental dataset.** Extent of LD variation observed in each continental dataset when haplotype block comprising the set of 18 variants identified in combined dataset were mapped to continental datasets.
